# Sensory Cues in Assistive Technologies: a Comparative Study of the BuzzClip and NOA

**DOI:** 10.1101/2025.02.20.25322389

**Authors:** Claire E. Pittet, Maël Fabien

## Abstract

Electronic travel aids have the potential to significantly improve the security and independence of blind or visually impaired people when traveling. However, it is critical that such devices convey relevant information without overwhelming users. This study examines the performance of two assistive technologies in obstacle detection: the BuzzClip, which uses ultrasonic sensors to provide haptic feedback, and NOA, which utilizes infrared cameras and an AI-powered computer to deliver audio feedback. One participant completed four different mobility routes with each route tested separately using both devices, and the number of auditory or haptic stimuli was recorded. According to the data collected, the BuzzClip produced significantly more sensory feedback than NOA. This difference can be attributed to the devices’ detection methods and the ability to filter obstacles. These findings highlight the differences between certain electronic travel aids, underlining the importance of creating tools that provide sufficient information to users while avoiding the potential sensory overload caused by excessive stimuli. Such considerations are crucial in designing mobility tools that effectively enhance mobility while offering an agreeable experience for users.

## Sensory Cues in Assistive Technologies: a Comparative Study of the BuzzClip and NOA

In recent years, many electronic travel aids (ETAs) have been developed to enhance safety and independence for blind or visually impaired (BVI) people when navigating. One of their primary objectives is to detect obstacles that cannot be detected by traditional mobility tools. For example, objects at upper body and head level can present risks for BVI individuals and are not detected with a white cane. Manduchi and Kurniawan (2010) found that within the more than 300 blind individuals they surveyed, at least one monthly head-level injury occurred for 13% of the respondents and 7% of them fell monthly while walking. These results demonstrate that although traditional mobility tools are essential in the life of BVI individuals, they have limitations.

Various approaches have therefore been explored to develop ETAs which expand spatial representation for BVI individuals. For instance, such devices often use ultrasonic sensors or cameras to gather information about the environment, and haptic and/or auditory feedback to convey information to the user (Dakopoulos & Bourbakis, 2010; Strumillo et al., 2018; Masal et al., 2023).

While these solutions offer distinct advantages, the critical design challenge is ensuring that the device provides sufficient and accurate obstacle information without overwhelming the user with excessive or complex cues. Indeed, too many cues or poor clarity can increase sensory load, interfere with other sensory inputs essential for navigation, complicate the required training, and increase mental fatigue when using the device (Kristjánsson et al., 2016; Yun & Deng, 2022; Roentgen et al., 2012; Elli et al., 2014).

This study focuses on the importance of balancing the amount of sensory feedback conveyed to avoid mental fatigue while still ensuring safety. To this end, we compare two devices which use different approaches: one uses ultrasonic sensors and haptic feedback, while the other utilizes infrared cameras and spatialized audio.

Our objective is to assess and compare the quantity of sensory cues from both solutions across different real-life mobility environments.

## Method

### Participants

The study included a single subject, a sighted woman aged 21-25, who participated blindfolded. The participant had a basic understanding of both devices used in the study. Prior to the assessment, she received training on the devices and the white cane, ensuring she could use them correctly during the trials. Since the objective of the study was to verify the devices’ sensory outputs and not the participant’s performance, using a sighted participant was deemed sufficient.

Informed written consent was obtained from the participant, and the research adhered to the principles of the Declaration of Helsinki. Ethical approval was not sought following a clarification of responsibility from the ethical committee (CER-VD).

## Materials

### BuzzClip

The BuzzClip is a compact device designed for obstacle detection that can be clipped onto clothing, handheld, or attached to a white cane, functioning similarly to a smart cane like the Ultracane (*https://ultracane.com/*) or WeWALK (https://wewalk.io/en/). It uses ultrasonic sensors to detect obstacles within a 30° field of view and provides haptic feedback to alert the user. The BuzzClip offers three detection ranges (1, 2, or 3 meters), with the 2-meter range used in this study (iMerciv, n.d.). For the trials, the device was attached to a white cane (see Fig. 1).

**Figure 1.**
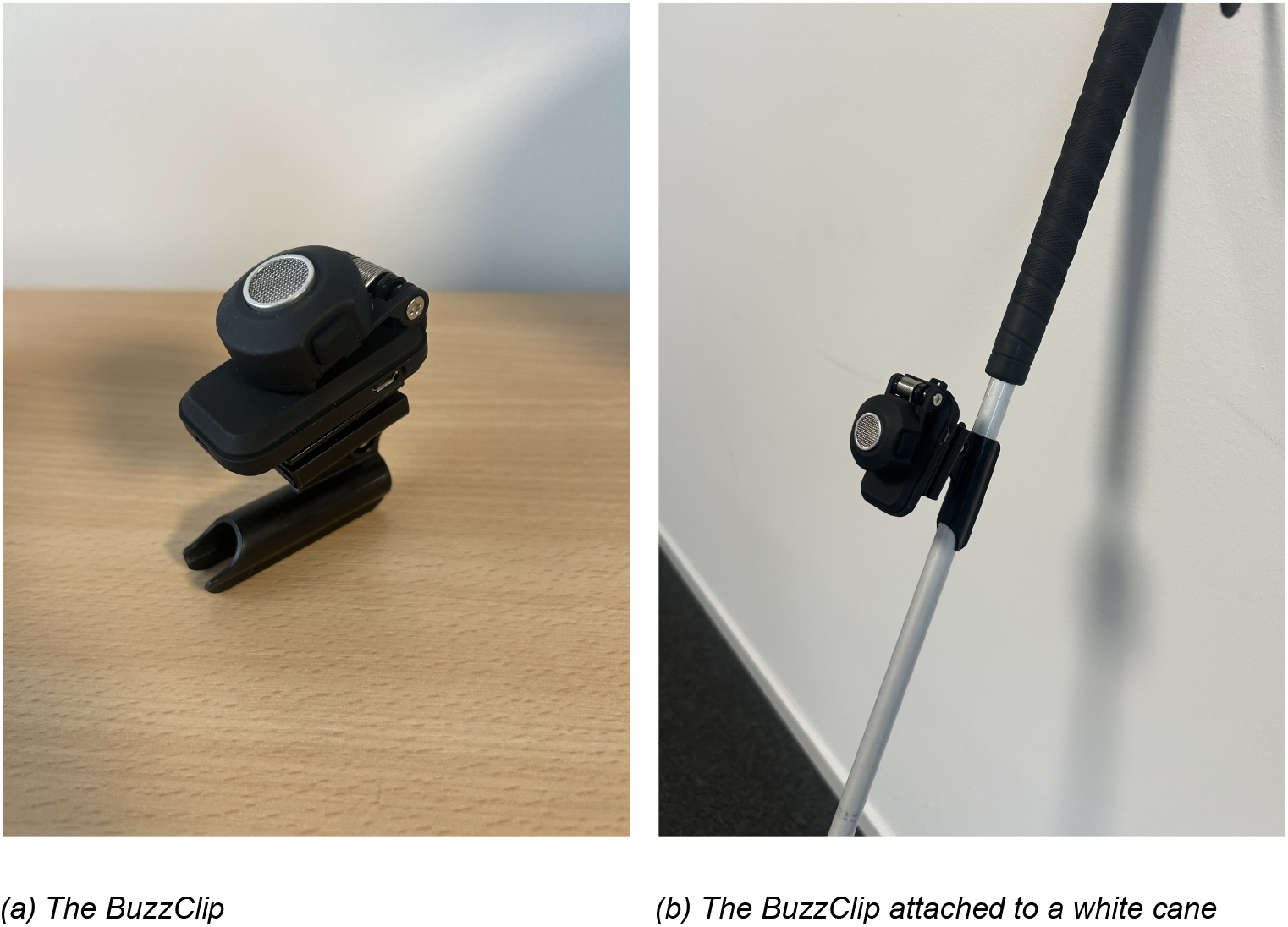
The BuzzClip Device

### NOA

NOA is a wearable mobility vest with an AI-based computer and wide-range infrared cameras designed for obstacle detection, GPS navigation, and AI object recognition (see Fig. 2). This study focuses exclusively on NOA’s obstacle detection feature. The cameras provide a field of view of 90° vertically and 170° horizontally, with a 0.3-10-meter range. NOA delivers 3D sound feedback, providing directional information on potential obstacles (https://biped.ai/). Like the BuzzClip, NOA was set to a 2-meter detection range for the trials.

**Figure 2.**
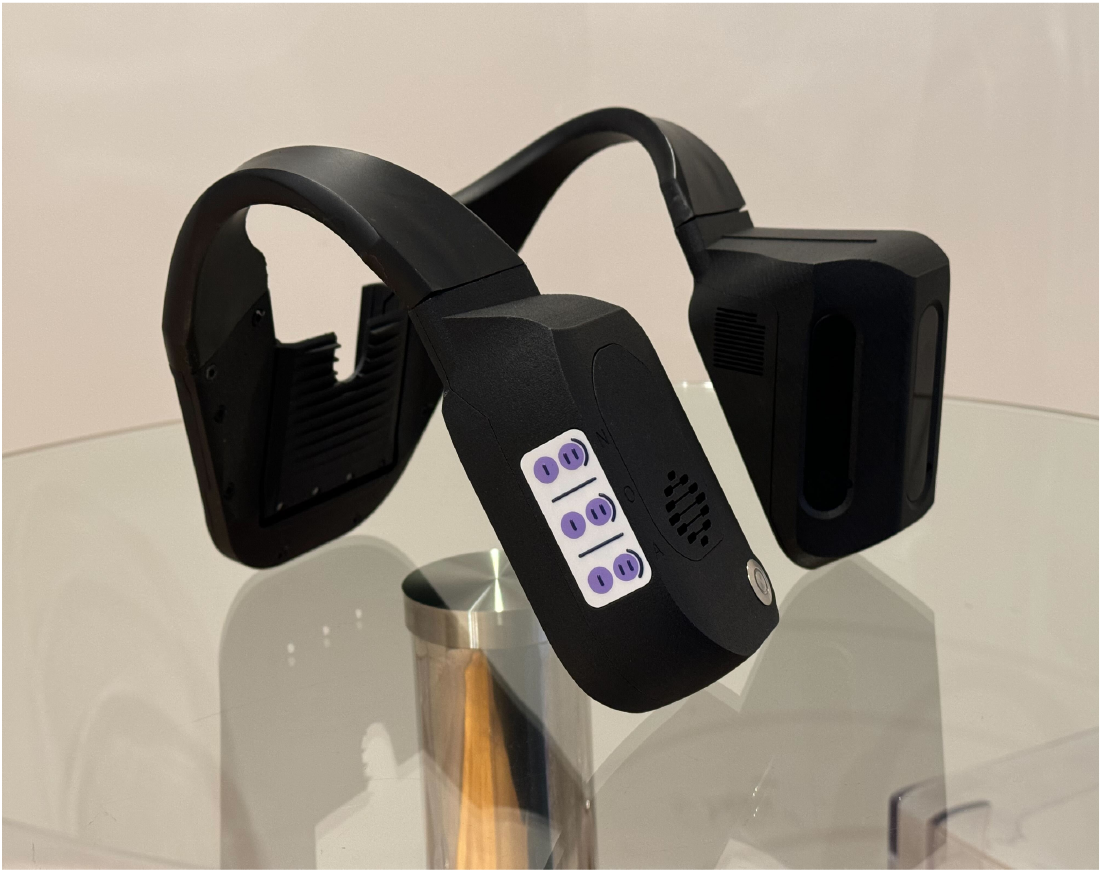
The NOA Device

## Procedure

Four routes were selected to represent various mobility scenarios. The blindfolded participant navigated each route twice: once using the NOA device with a white cane and once using the BuzzClip clipped onto the cane. Each trial documented the sensory cues from each device.

To ensure accurate data collection, each trial was filmed. This enabled the research team to review the footage and count the number of cues detected by each device, as well as understand the types of obstacles identified.

For the BuzzClip, the participant verbally indicated each time she felt a vibration. This was not required for NOA, as the auditory cues were played through a loudspeaker, making them audible on video. Each individual obstacle detected was considered one cue, regardless of the duration of the signal.

### Indoor Corridor

The first task was conducted in an indoor hallway, approximately 2 meters wide and 30 meters long (see Fig. 3). The corridor was linear, with no dynamic or static obstacles present during the task. The only detectable obstacles in the environment were the walls on both sides of the participant.

**Figure 3.**
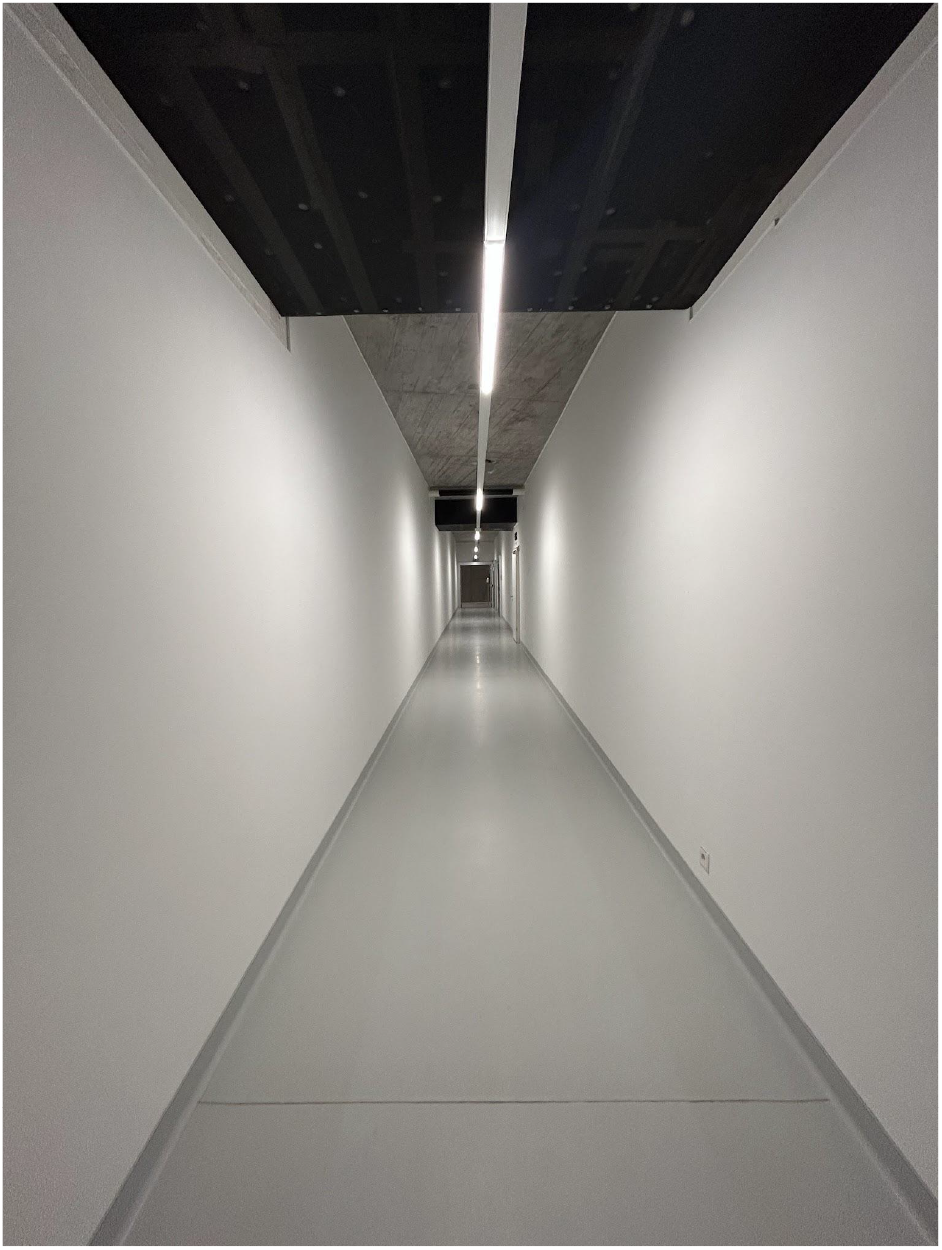
Indoor Corridor Course

### Construction Site

The second course was performed on a secure path through a construction site (see Fig. 4). The path spanned approximately 20 meters in length and 2.5 meters in width, with a slight uphill incline and a gentle turn at the midpoint. The path was free of pedestrians, and the only static landmarks were the barriers on either side of the path and a restaurant display board located at the endpoint.

**Figure 4.**
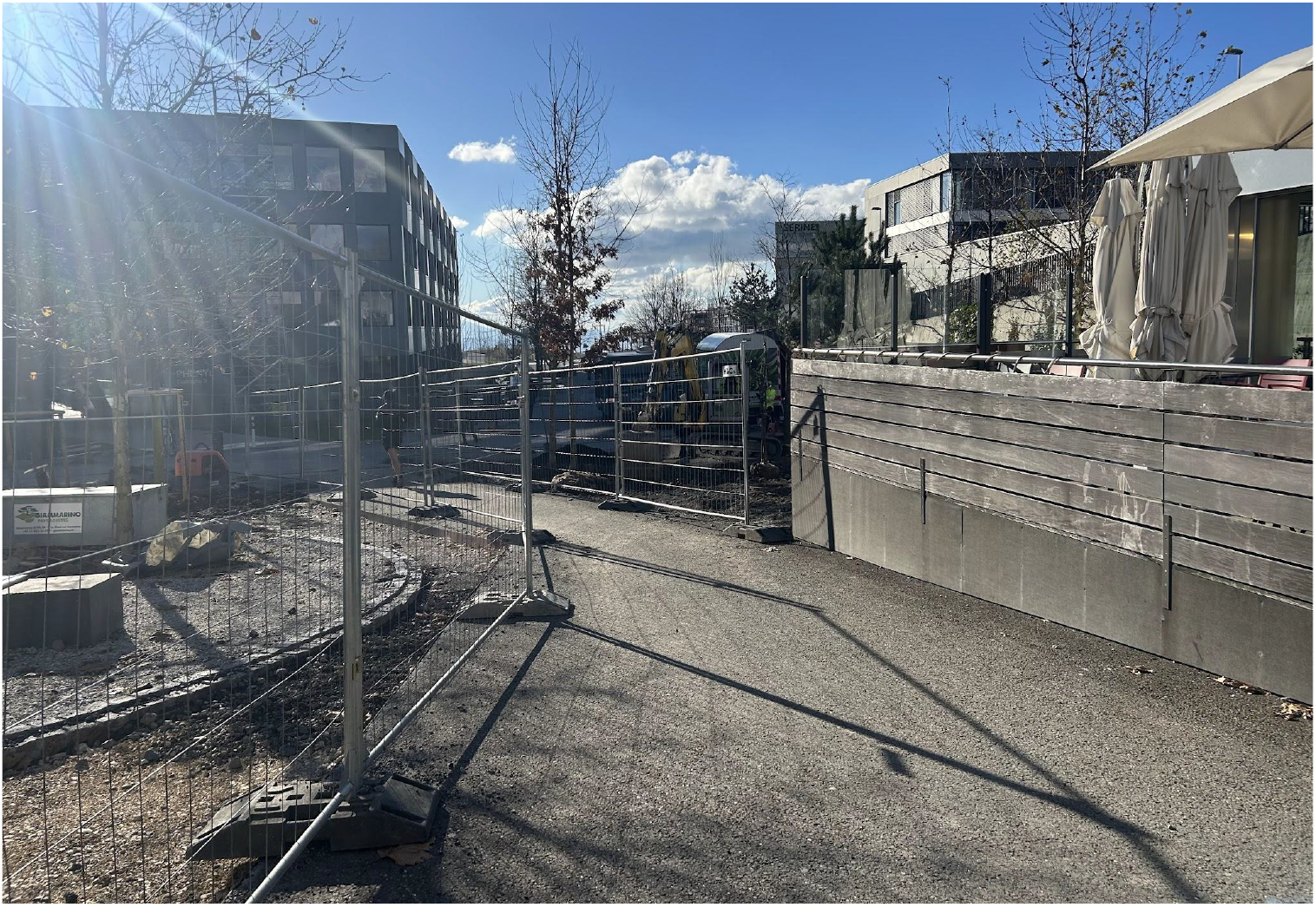
Construction Site Course

### Wooded Area

The third path was designed to simulate conditions commonly found in a park or forest environment (see Fig. 5). The straight path was approximately 15 meters in length and 1.5 meters in width. The surface consisted of wood chips, with weeds and small plants growing along the sides. Additionally, a small tree overhung the path, with branches extending at both ground and head height.

**Figure 5.**
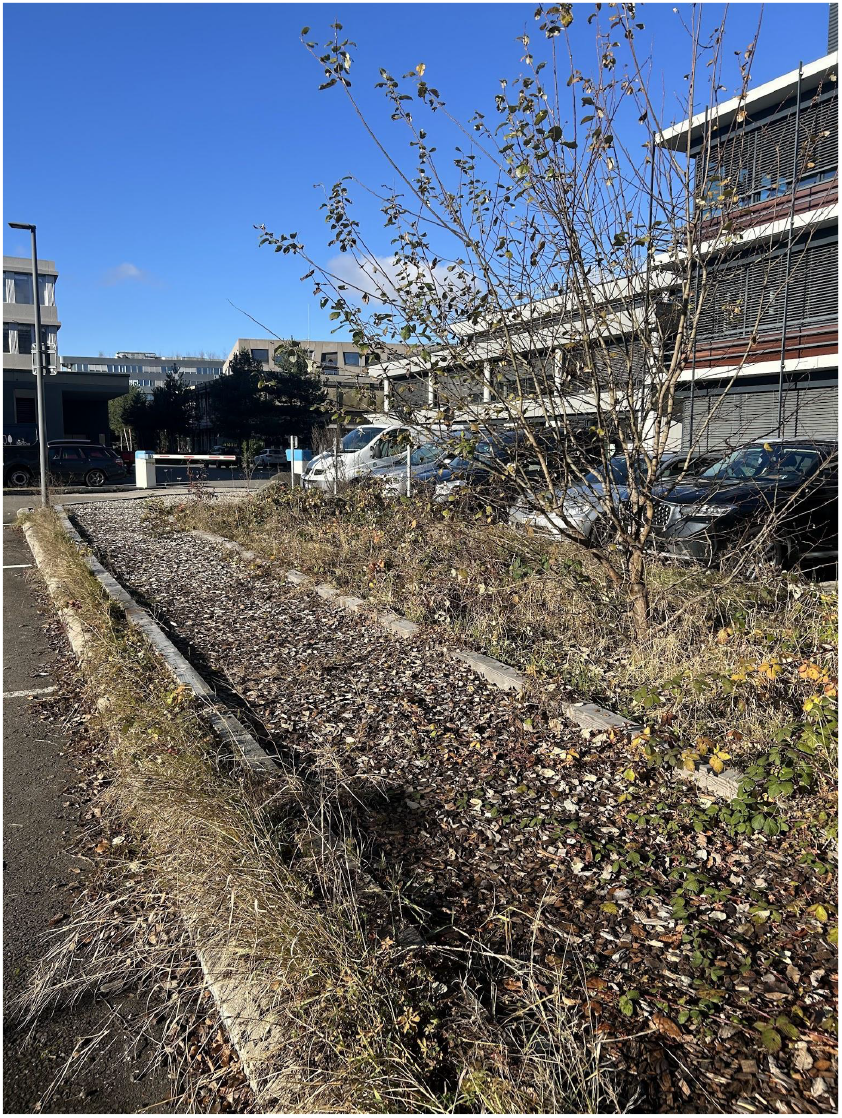
Wooded Area Course

### Residential Area

The final course was conducted on a quiet residential street (see Fig. 6). The path measured approximately 25 meters in length, with a sidewalk about 2 meters wide. On the left side, parked cars were present, while the right side featured an open grassy area with two trees, garbage bins, and a lamp post at various intervals along the route. No pedestrians or additional obstacles were present on the sidewalk during either of the two trials.

**Figure 6.**
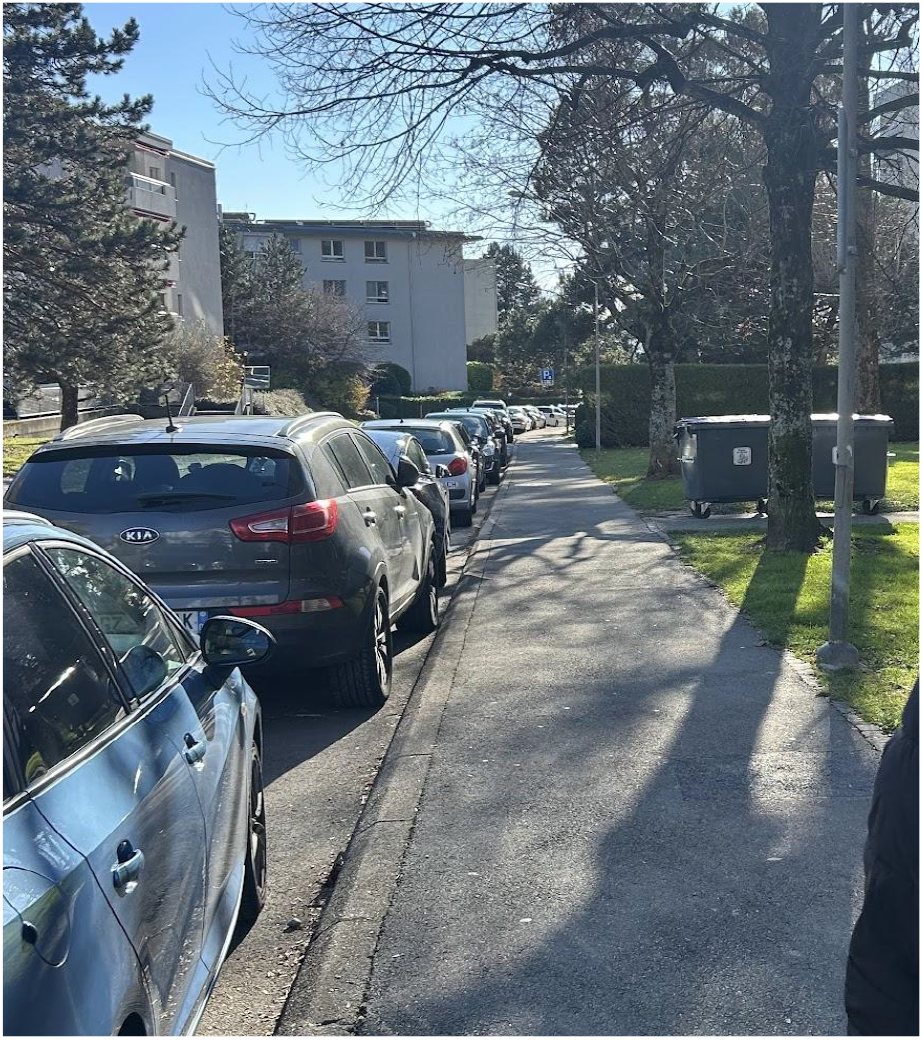
Residential Area Course

## Results

The results from the trials are presented for each of the four courses: Indoor Corridor, Construction Site, Wooded Area, and Residential Area. The number of cues detected by each device, NOA and BuzzClip, is shown in Figure 7.

**Figure 7.**
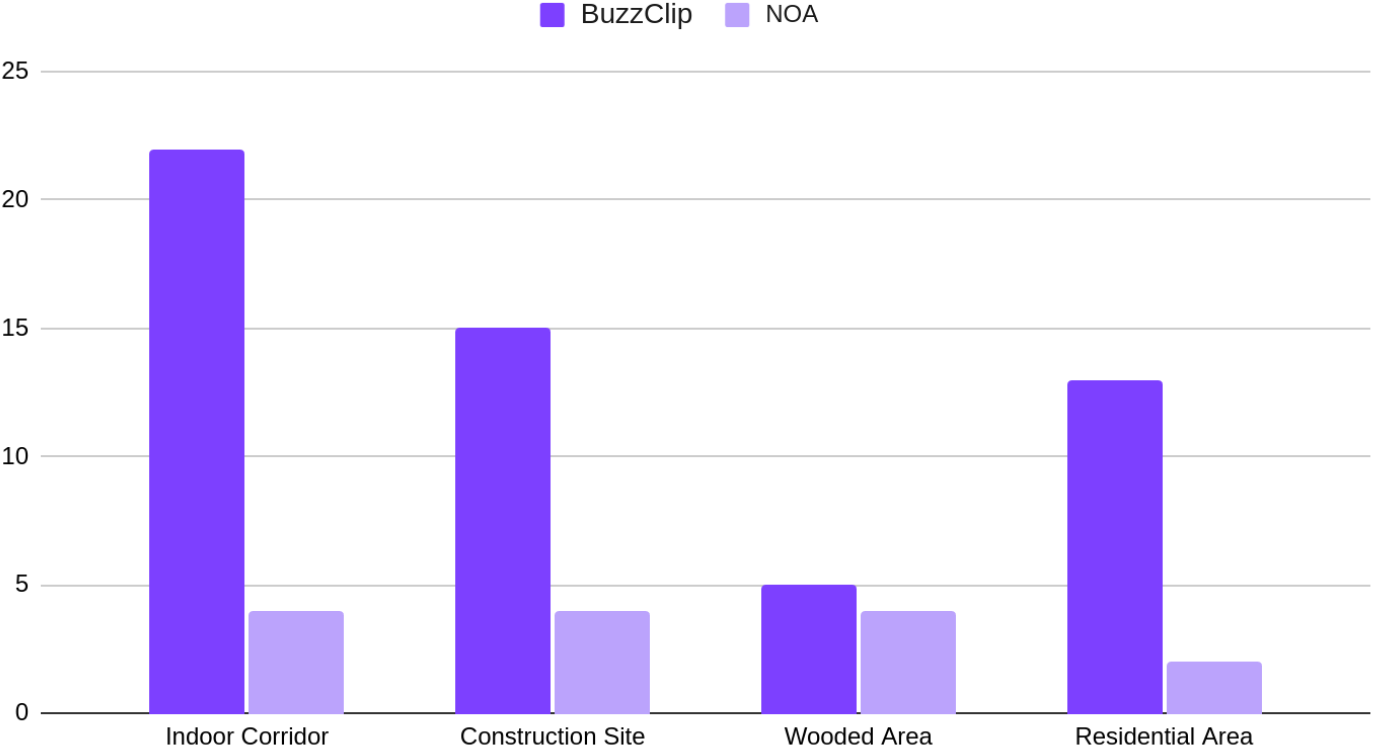
Comparison of Obstacles Detected by the BuzzClip and NOA Across Different Routes

In the indoor corridor, the BuzzClip detected 22 obstacles, while NOA registered only 4. Based on the footage, the BuzzClip signaled the presence of most walls due to the cane’s sweeping motion. In contrast, NOA only issued warnings when the participant deviated from the wall-trailing path.

Similar results were observed on the construction site path, where the BuzzClip detected 2.75 times more cues than NOA. Specifically, the BuzzClip detected 15 obstacles along the 20-meter path, primarily triggered by the cane’s motion, with only one actual risk of collision when the participant deviated from the path. In contrast, NOA issued only 4 warnings, all linked to potential collisions with obstacles.

On the wooded path, similar results were observed with both devices (NOA = 4, BuzzClip = 5). Both devices detected the weeds and plants along the sides, as well as the overhanging tree in the middle of the course.

In the residential area, the BuzzClip detected 13 obstacles, while NOA detected 2, yielding 5.5 times more cues from the BuzzClip. As in previous trials, the BuzzClip frequently identified side objects like cars and other landmarks, even when no collision risk was present.

## Discussion

The findings revealed that both devices’ performance varied significantly in the majority of the mobility scenarios. The BuzzClip consistently detected a higher number of obstacles, largely due to its use of ultrasonic sensors and its attachment to the cane, which caused it to sense objects on the sides and continuously activate during the sweeping motion. Although frequent obstacle detection has its benefits, users may experience mental fatigue, as frequent alerts require constant processing. This factor could potentially impact the overall efficiency of such devices, especially in complex environments.

Conversely, NOA’s obstacle detection was more selective, likely due to its AI-based filtering and shoulder-mounted position. This capability could potentially reduce cognitive load by minimizing unnecessary alerts.

One advantage of the study was that it was carried out in semi-controlled settings, which allowed for consistent comparisons of the devices in real-world situations. However, validity was significantly affected by the use of a single sighted blindfolded subject. Although the objective of the study was to evaluate the devices’ stimuli rather than their use by real users, future research should involve larger samples of BVI participants to enhance the generalizability of the results.

## Conclusion

The findings illustrate the strengths and limitations of ultrasonic devices like the BuzzClip, which provide high sensitivity but run the risk of overstimulation with frequent cues. Conversely, the selectivity of computer- and camera-based devices such as NOA, may reduce the mental load often problematic with ETAs. Future research should explore testing in additional environments, with more BVI participants and devices, and also collecting qualitative feedback to better understand user experiences.

## Data availability statement

The data that support the findings of this study are available from the corresponding author, C. E. Pittet, upon reasonable request.

